# Social return on investment of home exercise and community referral for people with early dementia

**DOI:** 10.1101/2021.06.23.21259333

**Authors:** Ned Hartfiel, John Gladman, Rowan Harwood, Rhiannon Tudor Edwards

## Abstract

**Background:** Exercise can improve physical function and slow the progression of dementia. However, uncertainty exists around the cost-effectiveness of exercise programmes for people with early dementia.

**Objective:** The aim is to determine whether a home-based supervised exercise programme (PrAISED – promoting activity, independence, and stability in early dementia) can generate a positive social return on investment (SROI).

**Methods:** SROI was conducted as part of a randomised controlled feasibility trial comparing PrAISED with usual care. Wellbeing valuation was used to compare the costs of the programme with the monetised benefits to participants, carers, and healthcare service providers.

**Results:** The PrAISED programme generated SROI ratios ranging from £3.46 to £5.94 for every £1 invested. Social value was created from improved physical activity, increased confidence, more social connection and PrAISED participants using healthcare services less often than usual care.

**Conclusion:** Home-based supervised exercise programmes can generate a positive SROI for people with early dementia.

**Trial registration:** ClinicalTrials.gov: NCT02874300 (first posted 22nd August 2016), ISRCTN: 10550694 (date assigned 31st August 2016)

## 1. Introduction

### 1.1 Dementia and physical activity

Dementia is a common and debilitating disease resulting in high costs to individuals and society. Public Health England has recognised dementia as a major priority. Studies show that delaying the onset of dementia by two years can have substantial economic and societal benefits (Rakesh et al., 2017). Research indicates that interventions that promote exercise and physical activity may slow the rate of dementia progression. A study from Finland reported that a twice-weekly, 12-month, supervised exercise programme at home significantly reduced the rate of decline in activities of daily living for people with dementia (Pitkala et al., 2013). This study reported reductions in hospital admissions and overall costs, suggesting that intensive exercise, with the right support, may be achievable and sustainable for improving the quality of life for people with dementia.

### 1.2 Promoting activity, independence and stability in early dementia (PrAISED)

PrAISED is a novel intervention in the UK to determine whether an individually tailored, home-based supervised exercise programme for people with early dementia can be clinically effective and cost-effective (Harwood et al., 2018). In alignment with World Health Organisation guidelines, the PrAISED exercise programme involves a minimum of 150 minutes of moderate to vigorous physical activity per week. In addition, the PrAISED programme involves referring people with early dementia to relevant community groups to encourage physical activity and social participation.

Between September 2016 and March 2018, a feasibility randomised controlled trial (RCT) of PrAISED was conducted at two sites, Derby and Nottingham. Patients with early dementia were randomised to a PrAISED programme consisting of moderate to high supervision (12 to 50 home sessions within twelve months), or to a usual care group consisting of a brief assessment of falls prevention (1 or 2 sessions within twelve months). Delivered by physiotherapists, occupational therapists and rehabilitation support workers, the PrAISED programme included assessment of patients, creation of individualised exercise plans, delivery of supervised exercises and activities, and referral to appropriate community activities. To encourage good habit-formation and the continuation of self-directed exercise, home visits were tapered over the twelve-month study period with more frequent sessions during the first three months.

Patients with early dementia were assessed at baseline and twelve months. The primary outcome measure was activities of daily living (ADLs) measured by the Disability Assessment for Dementia (DAD) scale. Secondary outcomes included physical activity, balance, mobility, fear of falling, quality of life, carer strain, and health service use. Additional methodological details are found in the study protocol (Harwood et al., 2018) and the feasibility trial findings relating to effectiveness found elsewhere (Goldberg et al., 2019). A social return on investment (SROI) analysis of the PrAISED feasibility trial was undertaken to help assess cost-effectiveness and potential value for money.

## 2. Methods

SROI is a pragmatic form of Social Cost Benefit Analysis (Social CBA), which is recommended in the HM Treasury Green Book to assess interventions that impact social welfare (New Economics Foundation, 2012). Using quantitative and qualitative methods, Social CBA measures and values all relevant costs and outcomes (HM Treasury, 2018). First documented in the early 2000s, SROI methodology has been refined and described in the Cabinet Office ‘A Guide to Social Return on Investment’ (Cabinet Office, 2012). SROI considers what outcomes are relevant and significant to stakeholders and then assigns financial proxies to outcomes which often do not have market values enabling findings to be reported in a common metric (£s).

Using a benefit-cost ratio comparing the value of outcomes with the value of inputs, the reporting of findings is easy to interpret and understandable to a wide audience. With the introduction of the Public Services (Social Value) Act 2012, the UK government is increasingly interested in evaluation methods which capture the social, economic and environmental outcomes of health and social care interventions. In taking a societal perspective, SROI considers the costs and benefits to key stakeholders. A societal approach is useful for informing decision-making within the NHS, which has budgets to be allocated between various programmes.

In this paper, SROI methodology was used to generate a range of SROI ratios using quantitative and qualitative data collected during the PrAISED feasibility trial. SROI methodology involves the following stages: identifying stakeholders, developing a theory of change, evidencing outcomes, valuing outcomes, calculating costs and estimating the SROI ratio. The SROI analysis in this study was conducted in accordance with the 21-item SROI Quality Assessment Framework Tool (Hutchinson et al., 2018) (Appendix 1).

### 2.1 Identifying stakeholders

The first stage was to identify the key stakeholders, which are the people or organisations significantly affected by the PrAISED programme. The PrAISED feasibility study research team, which included patient and public involvement representatives, identified three key stakeholders: patient participants, carer participants and the NHS. It was expected that patient participants would benefit from PrAISED exercises and community referrals; carer participants would benefit from the additional support provided by home visits from PTs, OTs and RSWs; and the NHS would benefit from less frequent health service use from patients participating in the PrAISED programme.

### 2.2 Developing a Theory of Change

A Theory of Change (ToC) model was created to identify the expected benefits experienced by key stakeholders. The ToC model illustrates the links between the inputs, outputs, outcomes and impacts of PrAISED (Figure 1).

**Figure 1:**
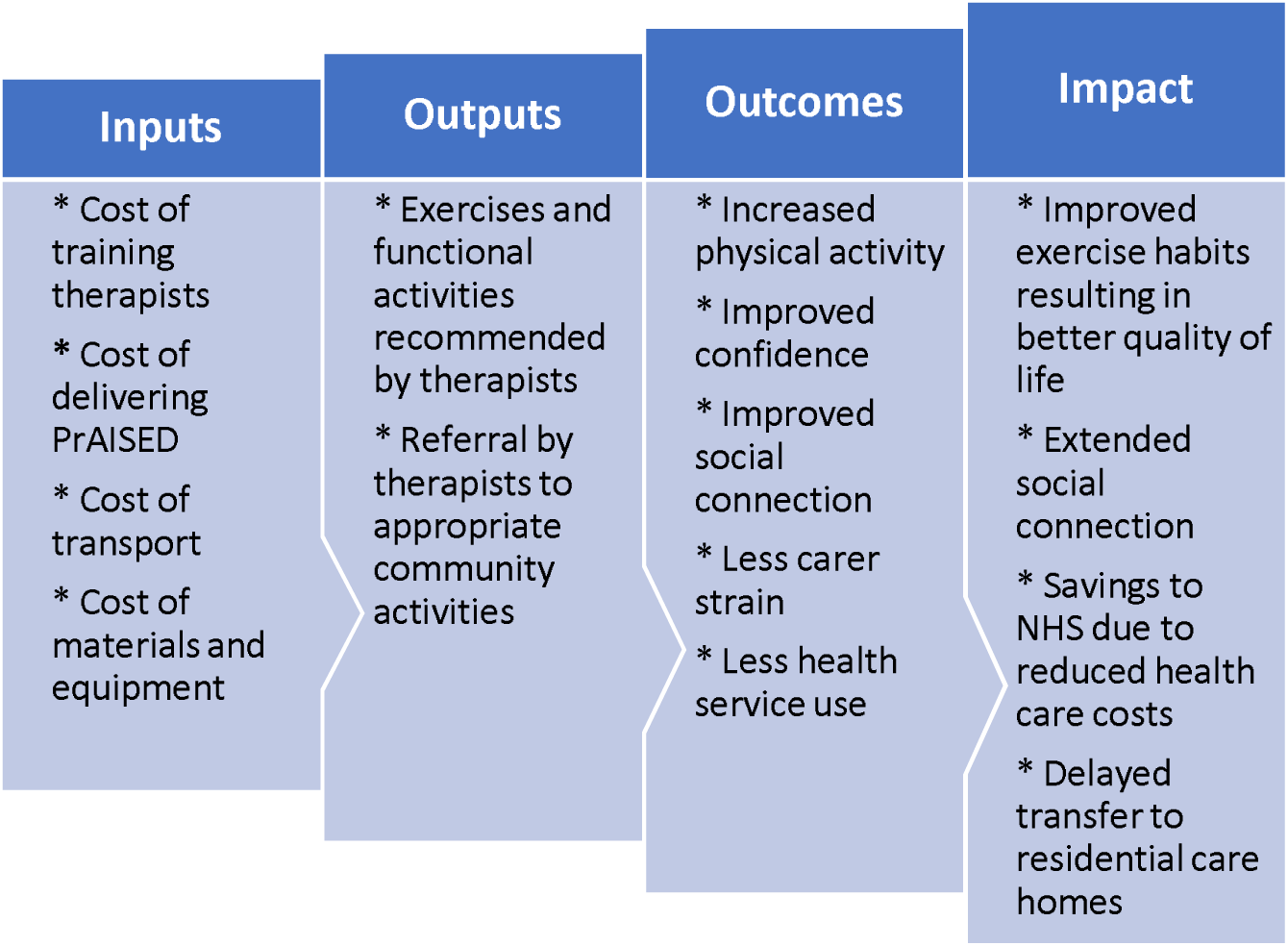
Theory of Change model.

### 2.3 Evidencing outcomes

After written informed consent was obtained from all participants, qualitative and quantitative data was collected to determine the effectiveness of PrAISED. Interviews were held with patients and carers after the twelve-month study. In addition, patients and carers completed baseline and 12-month questionnaires, which measured activities of daily living (ADL), physical activity, balance, mobility, fear of falling, quality of life, carer strain, and health care service use. Evidencing outcomes involved determining the amount of benefit experienced by the three key stakeholder groups: patient participants, carer participants, and the NHS (Table 1)

**Table 1:** Outcome measures for three key stakeholders.

| Stakeholder | Outcome measure | Completed by | Outcome |
| --- | --- | --- | --- |
| Patient participant | Disability Assessment for Dementia Scale (DAD) | Carer participant | ADLs |
| Patient participant | International Physical Activity Questionnaire | Carer participant | Physical activity |
| Patient participant | Berg Balance Scale | Patient participant | Balance |
| Patient participant | Timed up and go (TUG) test | Patient participant | Balance and mobility |
| Patient participant | Falls Efficacy Scale – International (FES-I) | Carer participant | Fear of falling |
| Patient participant | Health-related quality of life (EQ5D-3L) | Patient participant | Health-related quality of life |
| Carer participant | Carer strain index (CSI) | Carer participant | Carer strain |
| NHS | CSRI form | Carer participant | Costs of health care resource use |

For patient participants and carer participants, two different outcome scenarios were estimated: a base case and a conservative case. The base case considered patient or carer participants who stayed the same or improved over the 12-month study period for a particular outcome. Because dementia is a progressive condition, no change in outcome over twelve months can be considered a positive result. The conservative case, however, counted only those patient or carer participants who improved for a particular outcome over 12 months.

### 2.4 Valuing outcomes

The next stage of SROI methodology was to determine the value of outcomes for each of the three key stakeholder groups. To monetise the outcomes for patient and carer participants, financial proxies were assigned from the Social Value Bank (SVB), which is a databank of methodologically consistent unit costs for outcome indicators (Trotter et al., 2014). SVB is based on ‘wellbeing valuation’, which is recognised in HM Treasury’s Green Book as a robust method for financial appraisal and evaluation.

Wellbeing valuation uses thousands of large UK national surveys to isolate specific variables and to determine the effect of those variables on a person’s wellbeing. Wellbeing valuation then establishes the equivalent amount of money needed to increase a person’s wellbeing by the same amount. In this study, wellbeing valuation was applied to quantify and monetise significant patient and carer participant outcomes for both the base case and conservative case scenarios.

### 2.5 Calculating costs

Total costs of the PrAISED programme included the following categories: training costs for therapists; transportation and staffing costs required for therapists to deliver the programme in patient homes, and equipment costs for instructional materials and exercise equipment.

### 2.6 Estimating the SROI ratio

SROI ratios for base case and conservative case scenarios were generated by dividing the social value per participant by the cost per participant. To avoid overestimating the SROI ratio, ‘deadweight’ (i.e., quantity of outcomes that would have happened anyway) is usually considered (Cabinet Office, 2012). However, due to the randomised controlled study design used in the feasibility trial, calculating deadweight was not necessary as the data collected from the usual care patient participants represented ‘what would have happened anyway’. Discounting was also not necessary in this evaluation as all benefits measured were within one year of the baseline assessment.

## 3. Results

After receiving NHS ethics approval, participant recruitment occurred between September 2016 and March 2017 (Goldberg et al., 2019). Dementia patients (n=60) and carers (n=54) were enlisted from memory assessment clinics, community health venues and online registers. The mean age of participants was 76 years for patients and 68 year for carers. Patient participants were mostly male (57%) and carer participants were mostly female (65%).

### 3.1 Valuing outcomes for patient and carer participants

Between September 2017 and March 2018, 19 PrAISED patients (with their carers) completed interviews (Appendix 2). Of the 60 patient participants, 82% (49/60) completed both baseline and twelve-month questionnaires, which made it possible to determine the percentage who improved, stayed the same, or worsened for each outcome measure. A significant outcome was based on a > 10% difference between the percentage of PrAISED and usual care participants who improved for each outcome at twelve-months. At twelve months, four significant outcomes were identified:

#### Outcome 1: Increased physical activity resulting in improved activities of daily living

At 12-months, the base case indicated that 43% (12/28) of PrAISED patient participants reported no deterioration or improved DAD scores compared with 21% (3/14) of usual care participants. The conservative case showed 25% (7/28) of PrAISED patient participants with improved DAD scores compared to 14% (2/14) of usual care participants (Table 2).

**Table 2:**
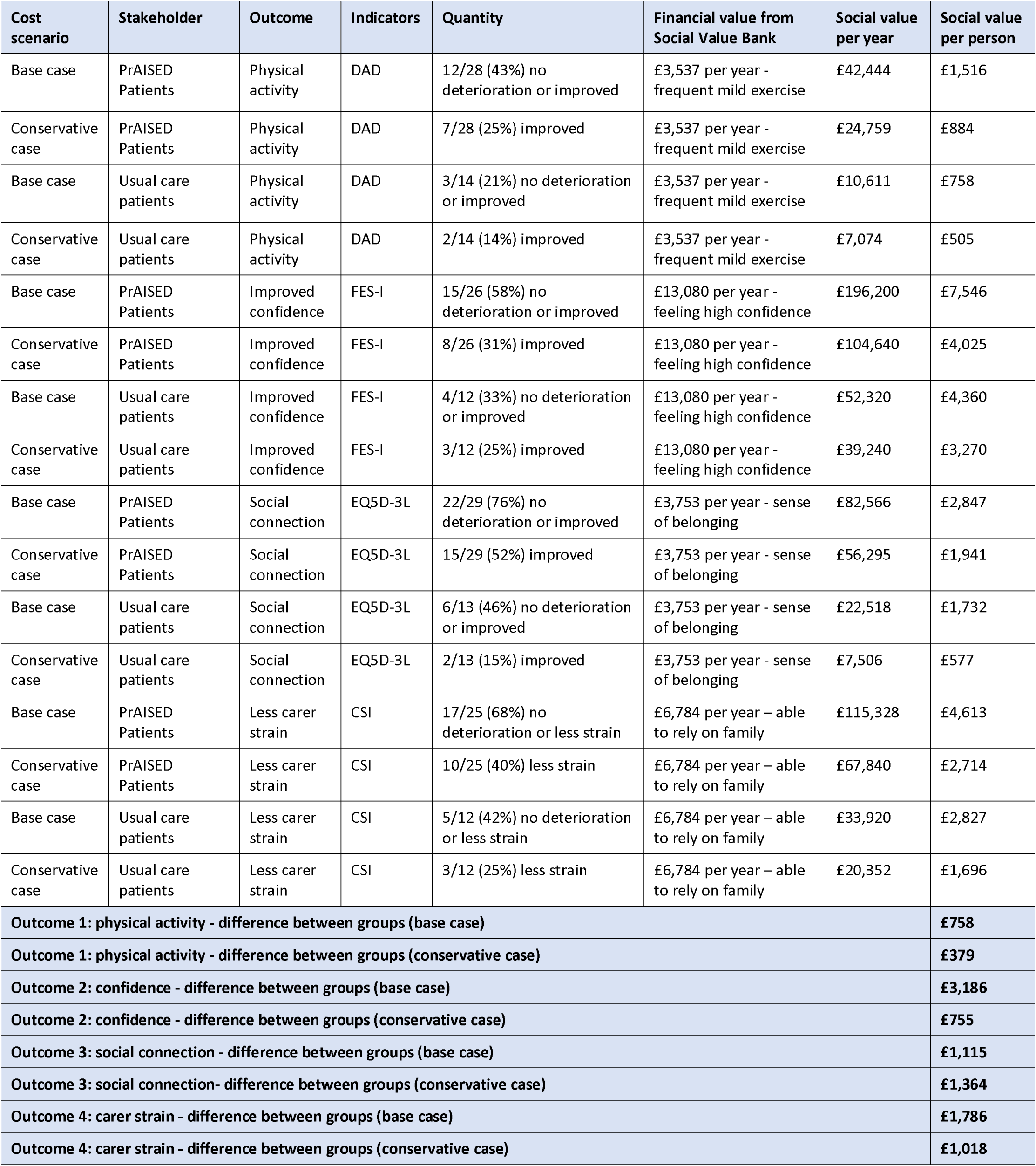
Valuing outcomes for patient and carer participants.

#### Outcome 2: More confidence resulting in less fear of falling

At 12-months, 58% (15/26) of PrAISED patient participants reported no change or less concern about falling compared with 33% (4/12) of usual care patient participants. The conservative case indicated 31% (8/26) of PrAISED patient participants with improved falls efficacy scores compared to 25% (3/12) of usual care participants (Table 2).

#### Outcome 3: More social connection resulting in improved quality of life

At 12-months, 76% (22/29) PrAISED patient participants reported no change or improved health-related quality of life compared with 46% (6/13) of usual care patient participants. The conservative case indicated 52% (15/29) of PrAISED patient participants with improved EQ5D scores at 12-months compared to 15% (2/13) of usual care patient participants (Table 2).

#### Outcome 4: Less carer strain (for carer participants)

At 12-months, 68% of PrAISED carer participants reported no deterioration or less carer strain compared to 42% of usual care participants. The conservative case indicated 40% (10/25) of PrAISED carers with reduced carer strain compared to 25% (3/12) of usual care carers (Table 2).

### 3.2 Valuing outcomes for the NHS

Outcomes for the NHS were assessed by measuring the health service use of patient participants during the 12-month study. Health service use for patient participants was completed by carer participants at baseline and 12 months via a Client Service Receipt Inventory (CSRI) form. Health service use included contact with GPs, nurses, outpatient services, accident and emergency (A&E) services, inpatient hospital days, physiotherapists and occupational therapy services. The 12-month results indicated that PrAISED patient participants used NHS health services less than the usual care patient participants by an average of £1,179 per person during the study (Table 3).

**Table 3:** Valuing outcomes for the NHS.

| Health service use<br>PrAISED patients (n=33)<br>Usual care patients (n=16) | Baseline | 12-mos | Quantity difference | Cost per visit | Total cost | Average cost per patient |
| --- | --- | --- | --- | --- | --- | --- |
| GP visits (PrAISED) | 48 | 38 | -10 | £37/visit<sup>1</sup> | -£370 | -£11.21 |
| GP visits (Usual care) | 18 | 23 | +5 | £37/visit<sup>1</sup> | +£185 | +£11.56 |
| Nurse visits (PrAISED) | 51 | 50 | -1 | £36/visit<sup>1</sup> | -£36 | -£1.09 |
| Nurse visits (Usual care) | 14 | 8 | -6 | £36/visit<sup>1</sup> | -£216 | -£13.50 |
| Outpatient services (PrAISED) | 51 | 23 | -28 | £125/visit<sup>2</sup> | -£3,500 | -£106.06 |
| Outpatient services (Usual care) | 28 | 12 | -16 | £125/visit<sup>2</sup> | -£2,000 | -£125.00 |
| A&E services (PrAISED) | 4 | 5 | +1 | £160/visit<sup>2</sup> | +£160 | +£4.85 |
| A&E services (Usual care) | 0 | 2 | +2 | £160/visit<sup>2</sup> | +£320 | +£20.00 |
| Inpatient hospital days (PrAISED) | 1 | 15 | +14 | £1,603/day<sup>2</sup> | £22,442 | +£680.06 |
| Inpatient hospital days (Usual care) | 0 | 18 | +18 | £1,603/day<sup>2</sup> | £28,854 | +£1,803.38 |
| PT/OT (PrAISED) | 6 | 4 | -2 | £44/visit<sup>1</sup> | -£88 | -£2.67 |
| PT/OT (Usual care) | 4 | 21 | +17 | £44/visit<sup>1</sup> | +£748 | +£46.75 |
| **Total (PrAISED)** |  |  |  |  |  | **£564** |
| **Total (Usual care)** |  |  |  |  |  | **£1,743** |
| **Difference between groups** |  |  |  |  |  | **£1,179** |

<sup>1</sup>Curtis & Burns, 2018, <sup>2</sup>NHS Improvement, 2018

### 3.3 Valuing inputs

The NHS was responsible for the training and delivery costs for the 12-month PrAISED programme. The equivalent of one full-time therapist (PT/OT) and four full-time RSWs delivered the PrAISED programme to 33 patient participants over the 12-month study period. The therapist and RSWs received two full days of PrAISED training prior to the start of the feasibility study. Training costs were estimated at £583 per therapist/RSW (Appendix 3). The difference in training and delivery costs between PrAISED and usual care patient participants was estimated at £1,351 (Appendix 3).

### 3.4 Calculating the SROI Ratio

The SROI ratio for the base case was £5.94 : £1, meaning that £5.94 of social value was generated for every £1 invested in the programme (Table 4). When the conservative case was considered, the total social value per participant was £4,672 compared with £8,024 in the base case, and the social value ratio was £3.46 of social value generated for every £1 invested in the programme (Table 4).

**Table 4:** SROI ratios.

| Cost scenario | Category | Difference between groups |
| --- | --- | --- |
| Base case | Outcome 1 – increased physical activity | £758 per participant |
| Conservative case | Outcome 1 – increased physical activity | £379 per person |
| Base case | Outcome 2 – improved confidence | £3,186 per participant |
| Conservative case | Outcome 2 – improved confidence | £755 per person |
| Base case | Outcome 3 – additional social connection | £1,115 per participant |
| Conservative case | Outcome 3 – additional social connection | £1,341 per person |
| Base case | Outcome 4 – less carer strain | £1,786 per participant |
| Conservative case | Outcome 4 – less carer strain | £1,018 per person |
|  | Outcomes for NHS | £1,179 per participant |
| **Base case** | **Total social value for all stakeholders** | **£8,024 per participant** |
| **Conservative case** | **Total social value for all stakeholders** | **£4,672 per person** |
| **Base case** | **Total cost** | **£1,351 per participant** |
| **Conservative case** | **Total cost** | **£1,351 per participant** |
| **Base case** | **SROI benefit/cost ratio** | **£5.94 : £1** |
| **Conservative case** | **SROI benefit/cost ratio** | **£3.46 : £1** |

## 4. Discussion

The results indicated that the PrAISED programme generated a positive SROI ratio ranging from £3.46 to £5.94 for every £1 invested. The findings showed that patient participants experienced social value through improved physical activity, confidence, and social connection, and carer participants through less carer strain. The results also revealed that PrAISED patient participants used NHS health services less often than usual care participants during the 12-month study.

### Comparison with other studies

The SROI ratios generated from the PrAISED intervention (£3.46 to £5.94: £1) were similar to SROI ratios generated in other related studies. For example, a study of peer support groups for people with dementia generated social value ratios ranging from £1.17 to £5.18 for every £1 invested (Willis et al., 2018); an evaluation of arts activities for people with dementia revealed social value ratios ranging from £3.20 to £6.62 for every £1 invested (Jones et al., 2020a); and a physical activity intervention for older people with chronic health conditions showed SROI ratios ranging from £2.60 to £5.16 for every £1 invested (Jones et al., 2020b).

### Strengths of this study

Although previous SROI evaluations have investigated the social value of arts activities and peer support groups for people with dementia, this is the first study to estimate the social value of physical activity and community referral to people with early dementia and their carer. A strength of this evaluation is the societal perspective and the inclusion of multiple stakeholder groups. Second, the results were strengthened by the randomised controlled study design, which is rare in SROI evaluations. Third, the validity of the results was strengthened from both quantitative and qualitative data collected from baseline and follow-up questionnaires completed by patients and their carer. Finally, the social value ratios calculated in this study were generated from value sets derived from wellbeing valuation, a consistent and robust method recommended in HM Treasury’s Green Book (2018) for measuring social CBA.

### Limitations of this study

First, there are only a limited number of pre-determined outcome values in the SVB derived from wellbeing valuation (Social Value UK, 2015). Not all outcome values in the SVB fit perfectly with the outcomes measured in health-related interventions. For example, high confidence in the SVB is valued at £13,080 per person per year. In our study, high confidence was derived from improvement in the Falls Efficacy Scale and applied to those patient participants who became more confident in walking and doing daily activities. SVB values, however, are typically binary, meaning that they are either applied in full or not at all. SVB valuations do not allow for situations where there may be varying degrees of improvement (Social Value UK, 2015).

Second, a common issue is that social value researchers working with the same data may arrive at different SROI ratios (Fujiwara, 2015). More social connection, for example, could be matched in the SVB with either ‘feeling belonging to neighbourhood’ (£3,753 per person per year) or ‘member of a social group’ (£1,850 per person per year). Matching outcomes from the study data with the most appropriate SVB value depends on the researcher’s discretion. This can introduce potential researcher bias and the possibility that social value estimates are upward-biased (Fujiwara, 2015).

Finally, the PrAISED programme was individualised. Although many of the exercises prescribed were the same, each patient received a personalised programme involving home exercises and referral to relevant community activities. Due to the specific skills of therapists and RSWs, and to the varied needs and abilities of patients, the individual PrAISED programmes differed considerably in the content delivered. For example, participant benefit could have been due to the PrAISED exercises or from community activities such as the parkrun, dementia-friendly swimming or ballroom dancing. The variety of activities prescribed makes it inappropriate to determine whether particular PrAISED activities or referrals were most responsible for participant outcomes.

### Further research

This SROI is based on quantitative and qualitative data from the PrAISED feasibility study with 60 patient participants and 54 carer participants. To confirm these results, a further SROI analysis will be required on the larger multi-centred randomised controlled trial of the PrAISED programme currently in progress (Bajwa et al., 2019). In addition, future SROI analysis could compare the social value ratios generated from both the face-to-face and online delivery of PrAISED.

## Data Availability

The availability of data referred to in this manuscript can be found here:
https://bmcgeriatr.biomedcentral.com/articles/10.1186/s12877-019-1379-5

### Appendix 1 SROI Quality Assessment Framework Tool

Appendix 1 – SROI Quality Assessment Framework Tool
| Research Question | Scoring | Notes |
| --- | --- | --- |
| 1) Was a well-defined question posed? | Yes, p.1 | The protocol for the PrAISED randomised feasibility trial included undertaking an SROI analysis (Harwood et al, 2018). |
| **Reasons for use of SROI method** |  |  |
| 2) Were authors transparent about why SROI methodology was chosen? (e.g. strategic planning/funding requirements) | Yes, p.1 | SROI methodology was chosen as part of a wider economic evaluation of the PrAISED programme (Harwood et al, 2018). |
| 3) Did authors report relevant background literature/justify the need for the study? | Yes, pp.1-2 | Relevant background information on the relevance of SROI analysis was provided at the start of the Methods section. |
| **Scope** |  |  |
| 4) Was the range of stakeholders included/excluded justified? | Yes, p.2 | The range of stakeholders and reasons for inclusion was provided in Section 2.1. |
| 5) Was the range of stakeholders wide enough to adequately answer the research question? | Yes, p.2 | The range of stakeholders was wide enough to adequately answer the research aim of estimating the SROI for the PrAISED programme. |
| 6) Was it clear how stakeholders were involved and what data would be gathered from them? | Yes, p.2 | The involvement of stakeholders, including PPI representatives, was provided in Section 2.1 and 2.3. |
| 7) Was ethics obtained/informed consent provided? | Yes, p.4 | Ethics approval was discussed at the start of Section 3. |
| **Theory of Change/Impact Map** |  |  |
| 8) Was the theory of change clear? i.e. the relationships between inputs, outputs and outcomes | Yes, p.2 | The Theory of Change model was presented and discussed in Section 2.2. |
| 9) Were unintended outcomes (positive/negative) detailed? | Yes, p.3 | Because dementia is a progressive condition, not all patients improved. This was discussed in Section 2.3. |
| **Study Design** |  |  |
| 10) Was the study design appropriate for the study question? (Control group, pre-post) | Yes, p.4 | The study design was appropriate for the study question. See Section 2.1. |
| 11) Was the sample described in detail/was the sample justified? | Yes, p.4 | The sample of 60 patient participants and 54 carer participants was sufficient for a feasibility trial. See Section 3. |
| **Analysis** |  |  |
| 12) Were inputs clear with non-monetized inputs valued appropriately? | Yes, p.6 | The inputs (i.e., training and delivery costs) were clearly presented in Section 3.3 and Appendix 3. |
| 13) Were capital costs, as well as operating costs included? | Yes, p.12 | Capital costs for equipment and stationery were presented in Appendix 3. |
| 14) Were costs that occur in the future 'discounted' to their present values? | Yes, p.3 | Discounting was not considered since all benefits were measured within one year of baseline measurements. See Section 2.6. |
| 15) Was dead-weight clearly described and calculated? | Yes, p.3 | Deadweight was not considered in this SROI evaluation due to the randomised controlled study design. See Section 2.6. |
| 16) Were the indicators valid and comprehensive? (Were the sources of all values clearly identified?) | Yes, p.5 | The indicators for significant outcomes and sources of financial value are clearly identified in Table 2. |
| 17) Were the proxies valid and comprehensive? (Were the sources of all values clearly identified?) | Yes, pp.3 -5 | To provide a consistent methodology, financial values were assigned from the Social Value Bank. See Table 2. |
| 18) Was length of benefit established and justified? (Drop-off) | Yes, p.4 | The length of benefit was established at 12 months in line with the duration of the intervention. See Section 3. |
| 19) Were limitations and biases reported? | Yes, p.8 | The limitations and biases were reported in Section 4. |
| 20) Was the final SROI ratio interpreted? | Yes, pp.7-8 | The range of final SROI ratios were presented and explained in Section 4. |
| 21) Was sensitivity analysis performed? Was justification provided for the range of values in the sensitivity analysis? | Yes, pp.3,6 | Sensitivity analysis presented base case and conservative case scenarios. See Sections 2.3 and 3.4. |

### Appendix 2 Selected quotes from interviews with patients and carers indicating physical, mental and social benefits of participation in PrAISED

#### Physical benefits

- ‘The physio exercises are important, because it’s keeping me fit’ (Patient participant, 1001).
- ‘My joints get a bit stiff these days…the exercises help to ease them.’ (Patient participant 1006)
- ‘With the help of the swimming (Alzheimer’s Group) and the other exercises, that’s given me more balance and strength.’ (Patient participant 1009)
- ‘Instead of doing the exercises, we’d go out for a walk. But the physio did suggest that, whilst you go out for your walks, you’re sort of implementing some of the exercises.’ (Patient participant 1011).
- ‘But that was the breakthrough, she took me out, she ran me up and down and said, you can run, do the park run. And that was it. And I’ve done thirty-nine now.’ (Patient participant 1017).
- ‘Yes, the programme has encouraged me to walk more and exercise more. I feel better for it. I walk to the shop and back again, trying to get the steps in.’ (Patient participant 1018)
- ‘You keep doing it and it works. It gets you back doing exercise. I used to play a lot of sport and got out of the habit, but it kept me going again.’ (Patient participant 1026)
- ‘It’s proved the benefits of those specific PrAISED exercises. His actual gait, when he walks, is a lot better than it was a year ago.’ (Carer participant for 1026)
- ‘The exercises made my back a lot better than it was.’ (Patient participant 2007)

#### Mental benefits

- ‘I feel great after all the exercises. Feel as though I’ve achieved something.’ It’s the activity from and the encouragement from PrAISED that has given me the confidence to go swimming (Alzheimer’s group). I’ve had positive encouragement from the group and quite a lot of improvements. The programme has given me a positive attitude.’ (Participant 1009)
- ‘It’s given me a boost, for a start, a very big boost.’ (Patient participant 1017)
- ‘I do different exercises. I go walking, I do the exercises and I do the gardening, and they’re all part and parcel. I was doing things that I never thought I could do.’ (Patient participant 1018).
- ‘It got you doing that life journey scrapbook, that’s something you wouldn’t have done. So then, Alzheimer’s had a carol concert, and they said would you give a talk at that? So you stood up with a mic in front of you, and you talked on living well with dementia.’ (Carer for patient participant 2012)

#### Social benefits

- ‘The people (therapist and RSW) who came were all very helpful and kind.’ (Patient participant 1017)
- ‘The parkrun has a social aspect to it. They’ve got to know me over there and chat to me and, it always pleases me because I come first in my age group.’ (Patient participant 1017)
- ‘SG (therapist or RSW) was here every week. And she loved walking. And she put me through my paces. And that was motivation for me, that I were going to beat her, I think I did a couple of times but not very often. And it’s, it’s embedded in me now, to carry on doing it. And I wish I could get her (therapist or RSW) to come again and do it for another year with me.’ (Patient participant 1018)
- ‘So he’s snooker on a Monday. Tuesday, you have been going to Man Cave. He started dementia-friendly swimming at the leisure centre. Thursday, we go to Healthy Hearts exercise class. Friday, you have been doing table tennis. And Sunday night, we’ve started ballroom dancing. So, that is how our life is changed because he found he could still do things.’ (Carer participant for 2012).

### Appendix 3 Supplementary tables for training and delivery costs

#### Training costs for two-day course (2 trainers, 1 therapist and 4 RSWs)

| Cost categories for training | Cost |
| --- | --- |
| Cost of two trainers (£195.17 per day per trainer x 2 trainers x 2 days) | **£781** |
| Cost of mileage for 7 people (2 trainers + 5 therapists) (25 miles average roundtrip x £0.45 per mile) | **£158** |
| Cost of manuals and printing for 5 therapists over 2-day training (£12.50 per person x 5) | **£63** |
| Cost of 1 therapist attending two-day training (£195.17 per day per therapist x 1 therapists x 2 days) | **£390** |
| Cost of 4 RSWs attending two-day training (£122.82 per day per RSW x 4 RSWs x 2 days) | **£983** |
| Cost of lunch and refreshments for 7 people (7 people x 2 days x £10 for lunch and refreshments) | **£140** |
| Cost of venue hire (£200 per day x 2 days) = £400 | **£400** |
| **Total Costs to deliver 2-day training for 7 people (2 trainers + 5 therapists)** | **£2,915** |
| **Average cost of a 2-day training per therapist/RSW** | **£583** |

#### Delivery costs for 60 patient participants

| Relevant information for calculating delivery costs | PrAISED | Usual care |
| --- | --- | --- |
| Number of participants with early dementia receiving PrAISED intervention | **33** | **16** |
| Average number of sessions delivered per participant | **23** | **2** |
| Total number of sessions delivered | **759** | **32** |
| Average time for therapist/RSW to deliver a session (including travel) | **2.5 hours** | **2.5 hours** |
| Total number of hours delivering PrAISED | **1,898 hours** | **80 hours** |
| Average number of sessions delivered per participant by registered therapists | **5 (22%)** | **1 session** |
| Average number of sessions delivered per participant by support workers | **18 (78%)** | **1 session** |
| Total number of registered therapist hours | **418 hours** | **40 hours** |
| Total number of support worker hours | **1,480 hours** | **40 hours** |
| Average mileage to and from participants' homes (round trip) | **25 miles** | **25 miles** |
| Total mileage to and from participants' homes | **18,975 miles** | **800 miles** |
| Total cost of mileage to and from participants' homes (£0.45 per mile) | **£8,539** | **£360** |
| Total cost of registered therapists (£26.92 / hour) | **£11,253** | **£1,077** |
| Total cost of support workers (£16.94 / hour) | **£25,071** | **£678** |
| Equipment Costs (£30 per participant) | **£990** | **£0** |
| Stationery costs (£5 per participant) | **£165** | **£0** |
| Training costs for one therapist and four RSWs | **£2,915** | **£0** |
| **TOTAL COSTS** | **£48,933** | **£2,115** |
| **TOTAL COSTS PER PARTICIPANT** | **£1,483** | **£132** |
| **DIFFERENCE IN TOTAL COSTS PER PARTICIPANT BETWEEN GROUPS** | **£1,351** |  |

